# Vegetarianism and mental health: longitudinal evidence in the 1970 British Cohort Study

**DOI:** 10.1101/2022.10.11.22280579

**Authors:** Thierry Gagné, Vanessa Kurdi

## Abstract

**Background:** Reducing animal product consumption has benefits for population health and the environment. The relationship between vegetarianism and mental health, however, remains poorly understood. This study explores this relationship in a nationally representative birth cohort in Great Britain.

**Methods:** We use data from the 1970 British Cohort Study, which collected information on diet at age 30 (*n* = 11,204) and psychological distress (PD) using the nine-item Malaise Inventory at ages 26, 30, 34, 42, and 46-48. We first developed a statistical adjustment strategy by regressing PD at age 30 on vegetarianism and 14 potential confounders measured at ages 10 and 26 (including PD at age 26). We then ran multilevel growth curve models, testing whether within-person changes in PD between ages 30 and 46-48 differed by vegetarianism, before and after statistical adjustment. Models were reproduced using red meat consumption at age 30 as a sensitivity analysis.

**Results:** At age 30, 4.5% of participants reported being vegetarian. In the cross-sectional models at age 30, vegetarians reported more distress compared with non-vegetarians in bivariate analysis (b = 0.30, 95%CI 0.09, 0.52), but this difference disappeared in the fully-adjusted model (b = 0.02, 95%CI -0.17, 0.21). In the longitudinal models between ages 30 and 46/48, there were no differences in within-person changes in psychological distress between vegetarians and non-vegetarians (*p* = .723). Sensitivity analyses yielded similar findings.

**Conclusion:** In this British cohort, vegetarianism at age 30 was not associated with changes in psychological distress during mid-adulthood. Since psychological distress in early adulthood predicted vegetarianism at age 30, more studies are needed to disentangle the progression of this relationship over the life-course.

## 1. INTRODUCTION

Reducing animal product consumption has clear benefits for population health and the environment.^1–4^ The relationship between vegetarianism and mental health is likely more nuanced. From a nutritional perspective, vegetarian diets may lead to mental health problems such as depression if they are not well-balanced in terms of nutrients, e.g., vitamin D and fatty Omega-3 acids, naturally found in animals such as fish or added to milk.^5^ From a psycho-social perspective, vegetarianism may lead to psychological distress in keeping with one’s capacity to act in line with their ethical or health values.^6,7^ Deviation from social norms regarding diet and consumption may also lead to feelings of marginalisation.^6,7^

Many studies suggest a negative association between vegetarianism and mental health.^8,9^ While 15-20 years have passed since studies first started exploring this relationship,^10,11^ the methodological quality of the current evidence remains surprisingly poor.^8,12–15^ One key issue concerns the reliance on weak study designs and its impact on the capacity to make causal statements. Importantly, the majority of studies linking vegetarianism and mental health are cross-sectional,^10,11,16–21^ making estimation inherently liable to issues of unobserved confounding (i.e., is the association explained by a third variable?) and temporality (i.e., if the association is causal, is it vegetarianism that increases the risk of mental health problems or it is poor mental health that increases the probability of starting vegetarianism over time?). For instance, a study in Germany retrospectively asked participants about the age of onset of vegetarianism and mental health problems and found that whereas these were associated, the timing at which participants started a vegetarian diet was likely to follow the incidence of mental health problems, instead of the other way around.^19^

New longitudinal studies are challenging the findings that cross-sectional studies yielded so far. In England, Northstone et al. (2017) examined depressive symptoms in parents of the Avon Longitudinal Study of Parents and Children (ALSPAC) over a two-year period (1994/95 to 1996/97) and found that vegetarianism was associated with an increased risk of depressive symptoms among men. However, this association was no longer significant once confounders - including subjective health and anxiety at baseline - were taken into account.^22^ Two studies using German and Chinese student samples recruited between 2012-16 and followed over one year similarly found no substantive associations between vegetarianism and changes in subjective wellbeing and mental health problems at the endpoint.^23,24^ One last Taiwanese study followed the incidence of depression in a large sample from 2005 to 2014 and found that vegetarians were 30% less likely to develop depressive symptoms during this period.^25^ However, since the prevalence of vegetarianism in China and Taiwan is high, the extent to which these findings may be generalised to countries with different food and social environments remains unclear.^4,16^

Robust evidence on the relationship between vegetarianism and mental health is critical to developing clear communication strategies and understanding windows of opportunity for population-level interventions promoting the reduction of animal product consumption. If the associations found in weaker studies are spurious, the misperception that vegetarianism “causes” mental health problems may prevent many from making a positive dietary change. Given rising levels of mental health problems over time, public health agencies may also be less willing to promote reducing animal product consumption if it leads to a higher risk of mental health problems.^26^

The aim of this study is to explore the relationship between vegetarianism and mental health across adulthood, leveraging data from a nationally representative cohort in Great Britain which collected data on diet at age 30 and psychological distress at multiple points between ages 26 (1996) and 46/48 (2016/18). We note that using longitudinal data does not solve *ipso facto* concerns regarding causal inference if confounding is not properly addressed. Molendijk et al. (2018) performed a systematic review of prospective studies linking diet and mental health and found that whereas a high-quality diet, including vegetarian, was associated with a subsequent lower risk of depression, most effect sizes became non-significant when adjusting for baseline differences in mental health.^27^ Therefore, we 1) develop a statistical adjustment strategy and 2) test differences in mental health trajectories between non-vegetarians and vegetarians during adulthood.

## 2. METHODS

### 2.1. Data

We use data from the 1970 British Cohort Study, which recruited 17,196 individuals born during a single week in April 1970 across the UK and followed them again at ages 5, 10, 16, 26, 30, 34, 38, 42, and 46-48. New participants were included between ages 5 and 26 to address the evolving ethnic distribution of Great Britain in the 1980s and 1990s. Ineligibility over time included prison, military service, emigration out of the UK, long-term loss to follow-up, and death. Response rates among eligible cases were generally above 70% across waves and are detailed in Supplementary Table 1.

A total of 11,216 participants responded in the age 30 wave in 2000. When exploring the confounders of the association between vegetarianism and psychological distress at age 30, we used the subset of 7,123 (63.3%) cohort members who also participated at ages 10 and 26. To test the relationship between vegetarianism and psychological distress, we used the subset of 6,712 (59.8%) cohort members who also participated at least once between ages 34 to 46-48.

### 2.2 Measures

#### Vegetarianism

Vegetarianism was measured at age 30 with the single question “Are you a vegetarian?” (Yes / No). Missingness was low (0.5%). This variable has been used elsewhere to test the relationship between intelligence in childhood and vegetarianism in adulthood.^28^

#### Psychological distress

Psychological distress was measured using two versions of the Malaise Inventory: a longer version with 21 items (ages 26, 30, and 34) and a shorter version with 9 items (ages 42 and 46-48). We used the 9-item version to have a consistent measure across waves.^29^ Items were dichotomous (Yes / No) and asked: 1) Do you feel tired most of the time?; 2) Do you often feel miserable or depressed?; 3) Do you often get worried about things?; 4) Do you often get into a violent rage?; 5) Do you often suddenly become scared for no reason?; 6) Are you easily upset or irritated?; 7) Are you constantly keyed up and jittery?; 8) Does every little thing get on your nerves?; 9) Does your heart often race like mad? Summation scores were computed, ranging from 0 (not distressed) to 9 (fully distressed). Missingness within waves varied from 3.2% at age 26, 1.4% at age 30, 0.8% at age 34, 14.6% at age 42, to 9.4% at age 46-48 (the items were administered in a self-completed questionnaire in later waves). The Malaise Inventory has been used elsewhere to understand mental health over the life-course.^30,31^

#### Covariates

To develop a robust statistical adjustment strategy, we selected a number of covariates measured at ages 10 and 26 based on the literature and information available in the 1970 cohort dataset. Whereas participants were also surveyed at age 16, we did not consider variables in this wave as recruitment problems (in part due to teacher strikes at the time) resulted in a much smaller sample size.

Variables at age 10 included: 1) *sex* (Male / Female); 2) *intelligence*, measured as a standardised score with a mean of 100 and a standard deviation of 15;^28,32^ 3) *health limitations reported by the mother* (No medical condition / Has a condition, does not affect life / Has a condition, affects life); 4) *family social class*, derived by the data management team based on the occupation of parents (Professional, managerial, or technical / Skilled / Partly or unskilled / Not available); 5) *meat consumption*, measured based on the question “How often do you eat each of these foods … Meat” (Nearly every day / Quite often / Sometimes / Hardly ever); 6) *country of residence* (England / Wales / Scotland).

Variables at age 26 included: 7) *cohabitation* (Living with a spouse / Living with a partner / Living alone or with others); 8) *parenthood* (Living with children / No); 9) *economic activity* (Employed / Unemployed / Out of the labor force); 10) *educational attainment*, derived by the data management team at age 30 (No qualifications / Secondary education / Pre-university / Further or higher education); 11) *body mass index*, based on weight at age 26 and height at age 30 (Underweight / Normal weight / Overweight / Obese); 12) *self-rated health* (Fair or poor / Good / Excellent); 13) *life satisfaction* (continuous score from 0 to 10); 14) *psychological distress*, using the same summation score from 0 to 9 based on the Malaise Inventory.

### 2.3 Statistical analysis

We first describe sample characteristics by vegetarian status, reporting significant pairwise differences using Chi-Square tests for categorical variables and Student’s t-test for continuous variables. We then perform our main analytic strategy in two separate stages.

First, to examine which variables may predict vegetarianism at age 30 and explain differences in psychological distress at age 30 by vegetarian status, we regressed psychological distress at age 30 on vegetarianism and all 14 variables at ages 10 and 26, entered first separately and then together in a fully-adjusted model. We report these findings using betas from linear regression models with robust standard errors.

Second, to examine whether changes in psychological distress between ages 30 and 46/48 differ between non-vegetarians and vegetarians, we ran a multi-level growth curve model based on a random-intercept linear model.^33^ Finding a significant joint interaction test between vegetarianism and time (coded using dummy terms) would support a difference in within-person changes of psychological distress between non-vegetarians and vegetarians over time. The model was done in three steps, entering sequentially: 1) vegetarian status and time; 2) vegetarian status, time, and covariates highlighted as confounders in the first stage; and 3) vegetarian status, time, covariates, and the interaction between vegetarianism and time. From the 14 variables that could be selected, we considered as meaningful those that changed the magnitude of differences in psychological distress at age 30 by vegetarian status by at least 10%. To better interpret the results of interactions, we also report the average marginal mean scores of psychological distress predicted by the fully-adjusted random-intercept models.^34^

Analyses were performed in a “complete-case” approach in Stata 17.^35^ Missing data are reported in Supplementary Table 2. Missingness across covariates within waves was low except for intelligence (25.2%), health limitations reported by the mother (9.4%), and meat consumption (16.5%). To mitigate the role of item-level and person-level (i.e., non-response across waves) missingness, analyses were also done across 20 imputed datasets in the age 30 sample (*n* = 11,261) using multiple imputation by chained equations.^36,37^ We discuss findings based on the complete-case samples, but both methods are reported in tables and show similar findings. We use a threshold of .05 to interpret findings as statistically significant.

As a sensitivity analysis, we reproduced our analyses using red meat consumption as an alternative definition of vegetarianism. Red meat consumption was measured at age 30 asking “How often do you eat red meat like beef, lamb, or pork?”, with seven response options ranging from “Never” to “More than once per day”. We dichotomized the variable and recoded participants into meat eaters and never meat eaters (9.1% of participants). These results support the main findings and are presented in Supplementary Tables 3 and 4.

## 3. RESULTS

### 3.1. Sample characteristics

Table 1 presents the distribution of sample characteristics. Out of 11,204 participants with valid data on vegetarian status at age 30, 500 (4.5%) reported being vegetarian. When asked about the type of diet participants followed, 11 (2.2%) reported having a vegan diet, 309 (61.8%) reported being vegetarian and eating dairy products, 168 (33.6%) reported being vegetarian but eating poultry and/or fish, and 12 (2.4%) reporting being some other kind of vegetarian.

**TABLE 1.** Sample characteristics of the 1970 British Cohort Study (1970 to 2016/18).

|  | Pairwise |  | Multiple imputation |  |
| --- | --- | --- | --- | --- |
|  |  |  | *n* = 11,261 |  |
| Variables | Non-vegetarian | Vegetarian | Non-vegetarian<br>95.5% | Vegetarian<br>4.5% |
| Distress at age 30 (0-9) (mean) | 1.55 | 1.77 | 1.55 | 1.77 |
| Distress at age 34 (0-9) (mean) | 1.66 | 1.80 | 1.71 | 1.82 |
| Distress at age 42 (0-9) (mean) | 1.82 | 1.93 | 1.89 | 2.02 |
| Distress at age 46/48 (0-9) (mean) | 1.71 | 1.94 | 1.81 | 2.00 |
| **Age 10** |  |  |  |  |
| **Sex (%)** |  |  |  |  |
| Male | 49.6 | 25.6 | 49.7 | 25.6 |
| Female | 50.4 | 74.4 | 50.3 | 74.4 |
| **Intelligence (45-147) (mean)** | 101.0 | 106.1 | 100.9 | 106.0 |
| **Physical/mental limitations (%)** |  |  |  |  |
| No medical condition | 75.1 | 77.1 | 74.8 | 77.0 |
| Yes, does not affect life | 10.9 | 8.9 | 11.1 | 8.9 |
| Yes, affects life | 14.0 | 14.0 | 14.1 | 14.1 |
| **Family social class (%)** |  |  |  |  |
| Professional, managerial, technical | 15.6 | 12.4 | 15.7 | 12.4 |
| Skilled | 47.5 | 44.8 | 47.5 | 44.8 |
| Partly or unskilled | 27.8 | 35.5 | 27.7 | 35.4 |
| Not available | 9.1 | 7.3 | 9.1 | 7.4 |
| **Country (%)** |  |  |  |  |
| England | 83.9 | 90.7 | 83.9 | 90.6 |
| Wales | 6.3 | 4.4 | 6.3 | 4.5 |
| Scotland | 9.8 | 4.9 | 9.8 | 4.9 |
| **Meat consumption (%)** |  |  |  |  |
| Nearly every day | 41.3 | 37.8 | 41.3 | 37.5 |
| Quite often | 38.7 | 33.1 | 38.6 | 33.2 |
| Sometimes | 15.9 | 17.5 | 15.9 | 17.6 |
| Hardly ever | 4.2 | 11.7 | 4.2 | 11.7 |
| **Age 26** |  |  |  |  |
| **Cohabitation (%)** |  |  |  |  |
| Living alone/other | 41.3 | 49.5 | 41.5 | 49.3 |
| Living with partner | 27.3 | 31.5 | 27.3 | 31.2 |
| Living with spouse | 31.4 | 19.1 | 31.3 | 19.5 |
| **Living with children (%)** |  |  |  |  |
| Yes | 24.8 | 15.3 | 25.7 | 16.1 |
| No | 75.3 | 84.7 | 74.3 | 83.9 |
| **Economic activity (%)** |  |  |  |  |
| Employed | 83.2 | 78.4 | 82.2 | 77.6 |
| Unemployed | 3.7 | 5.4 | 4.3 | 5.8 |
| Out of labor force | 13.0 | 16.2 | 13.5 | 16.6 |
| **Education (%)** |  |  |  |  |
| No qualifications | 13.4 | 6.2 | 13.6 | 6.3 |
| Secondary education | 40.5 | 29.0 | 40.4 | 28.9 |
| Pre-university | 14.2 | 12.6 | 14.2 | 12.6 |
| Further/Higher education | 31.8 | 52.2 | 31.8 | 52.2 |
| **Body mass index (%)** |  |  |  |  |
| Underweight | 2.9 | 5.7 | 2.9 | 6.1 |
| Normal weight | 66.2 | 80.4 | 65.0 | 79.6 |
| Overweight | 23.9 | 10.4 | 24.7 | 10.6 |
| Obese | 7.1 | 3.5 | 7.4 | 3.8 |
| **Self-rated health (%)** |  |  |  |  |
| Fair or poor | 9.1 | 8.9 | 9.7 | 9.5 |
| Good | 55.0 | 59.5 | 55.2 | 59.6 |
| Excellent | 35.9 | 31.6 | 35.1 | 30.9 |
| **Life satisfaction (0-10) (mean)** | 6.32 | 6.34 | 6.26 | 6.30 |
| **Distress (0-9) (mean)** | 1.74 | 2.13 | 1.80 | 2.17 |

The range of continuous variables (e.g., 0-9) is shown in parentheses. MI = Multiple imputation.

Vegetarianism was significantly associated with the majority of covariates. Among variables measured at age 10, vegetarian status was more common in women (74.4% *versus* 50.4% in non-vegetarians), those from a partly skilled or unskilled family background (35.5% *versus* 27.8%), and those growing up in England (90.7% *versus* 83.9%). Vegetarianism was more common in those who hardly ever ate meat at age 10 (11.7% *versus* 4.2%), and was also associated with a higher mean intelligence score (106 *versus* 101). Vegetarianism was not associated with health limitations.

Among variables measured at ages 26/30, vegetarian status was more common in those who completed further or higher education (52.2% *versus* 31.8% in non-vegetarians), not living with children (15.3% *versus* 24.8%), not living with a spouse (19.1% *versus* 31.4%), not employed (78.4% *versus* 83.2%), and underweight (5.7% *versus* 2.9%). Vegetarian status was also associated with a higher psychological distress score (2.1 *versus* 1.7). It was, however, not associated with self-rated health and life satisfaction at age 26.

### 3.3. Explaining differences in psychological distress at age 30 by vegetarian status

Table 2 presents results from the “cross-sectional” linear models regressing psychological distress at age 30 on vegetarian status, controlling for the selected variables at ages 10 and 26.

**TABLE 2.** Differences in psychological distress by <u>vegetarian status</u> at age 30 in the 1970 British Cohort Study.

|  | Complete-case sample |  |  | Multiple imputation |  |  |
| --- | --- | --- | --- | --- | --- | --- |
|  | *n* = 4,658 |  |  | *n* = 11,261 |  |  |
|  | B | 95%CI | % reduced | B | 95%CI | % reduced |
| **Vegetarianism at age 30 (ref. = No)** | **0.302** | **0.087, 0.516** | --- | **0.217** | **0.058, 0.377** | --- |
| **Controlling for ...** |  |  |  |  |  |  |
| **At age 10** |  |  |  |  |  |  |
| Sex | 0.200 | -0.013, 0.413 | 33.8 | 0.102 | -0.058, 0.263 | 53.0 |
| Intelligence | **0.358** | **0.146, 0.570** | -18.5 | **0.291** | **0.132, 0.451** | -34.1 |
| Health limitations | **0.294** | **0.079, 0.508** | 2.6 | **0.222** | **0.062, 0.382** | -2.3 |
| Family social class | **0.318** | **0.105, 0.532** | -5.3 | **0.236** | **0.077, 0.395** | -8.8 |
| Meat consumption | **0.273** | **0.057, 0.490** | 9.6 | **0.195** | **0.034, 0.355** | 10.1 |
| Country | **0.304** | **0.089, 0.519** | -0.7 | **0.218** | **0.058, 0.378** | -0.5 |
| **At age 26** |  |  |  |  |  |  |
| Cohabitation | **0.286** | **0.072, 0.501** | 5.3 | **0.204** | **0.044, 0.365** | 6.0 |
| Living with children | **0.321** | **0.107, 0.536** | -6.3 | **0.247** | **0.088, 0.406** | -13.8 |
| Economic activity | **0.278** | **0.062, 0.494** | 7.9 | **0.191** | **0.029, 0.353** | 12.0 |
| Education | **0.356** | **0.143, 0.570** | -17.9 | **0.295** | **0.136, 0.453** | -35.9 |
| Body mass index | **0.266** | **0.050, 0.482** | 11.9 | **0.190** | **0.029, 0.350** | 12.4 |
| Self-rated health | **0.267** | **0.056, 0.479** | 11.6 | **0.198** | **0.040, 0.356** | 8.8 |
| Life satisfaction | **0.295** | **0.084, 0.507** | 2.3 | **0.222** | **0.063, 0.381** | -2.3 |
| Psychological distress | 0.079 | -0.109, 0.267 | 73.8 | 0.001 | -0.145, 0.147 | 99.5 |
| **All 14 variables at ages 10 and 26** | 0.022 | -0.168, 0.212 | 92.7 | -0.032 | -0.180, 0.116 | > 100.0 |

The outcome is psychological distress (0-9) at age 30, defined through the 9-item Malaise Inventory. A higher score indicates higher distress. Estimates represent mean differences in the outcome between vegetarians and non-vegetarians, controlling for other variables. The "% reduced" represents the attenuation by statistical adjustment and is computed from the ratio of estimates (e.g., $1 - (0.022/0.302) = 92.7\%$ ). Bolded estimates are significant at the $p < .05$ level. CI = Confidence interval.

In the bivariate model, vegetarian status was associated with a 0.302 (95%CI 0.087, 0.516) higher psychological distress score (compared to non-vegetarian status). As a reference point, the standard deviation for psychological distress at age 30 was 1.758, suggesting a relatively small effect size. This association (i.e., the beta representing the difference between groups) remained significant when controlling for the majority of variables except two: sex, which decreased the magnitude of the association by 33.8%, and psychological distress at age 26, which attenuated the magnitude of the association by 73.8%.

We note that controlling for intelligence at age 10 and educational attainment by age 30 increased the magnitude of differences in psychological distress between vegetarians and non-vegetarians by 18.5% and 17.9%, respectively. That is, since intelligence and education were associated with a higher probability of vegetarianism and a lower distress score at age 30, adjusting for them resulted in a stronger association between vegetarianism and psychological distress.

Including all variables reduced the effect size of vegetarianism to *b* = 0.022 (95%CI -0.168, 0.212), representing a 92.7% attenuation towards the null (i.e., 1 – (0.022 / 0.302)).

### 3.3. Differences in psychological distress between ages 30 and 46/48 by vegetarian status

Table 3 presents the results from the longitudinal, multilevel growth curve model regressing the repeated measurements of psychological distress at ages 30, 34, 42, and 46/48 on vegetarianism at age 30, controlling for variables highlighted as potential confounders (i.e., attenuation of cross-sectional differences at age 30 by 10% or more, see Table 2): sex, intelligence at age 10, body mass index, self-rated health, and psychological distress at age 26, and educational attainment by age 30.

**TABLE 3.** Differences in psychological distress by <u>vegetarian status</u> between ages 30 and 46/48. 1970 British Cohort Study (1970 to 2016/18).

|  | Complete-case sample |  |  |  |  |  | Multiple imputation (20 datasets) |  |  |  |  |  |
| --- | --- | --- | --- | --- | --- | --- | --- | --- | --- | --- | --- | --- |
|  | *N participants = 4,909, n obs. = 16,727* |  |  |  |  |  | *N participants = 11,261, n obs. = 45,044* |  |  |  |  |  |
|  | No controls |  | With controls |  | With interaction |  | No controls |  | With controls |  | With interaction |  |
|  | B | 95%CI | B | 95%CI | B | 95%CI | B | 95%CI | B | 95%CI | B | 95%CI |
| **Vegetarianism at age 30 (ref. = No)** | **0.266** | **0.074, 0.458** | 0.051 | -0.102, 0.204 | 0.100 | -0.103, 0.303 | **0.162** | **0.011, 0.313** | -0.017 | -0.141, 0.107 | 0.037 | -0.119, 0.196 |
| **Time** |  |  |  |  |  |  |  |  |  |  |  |  |
| Age 30 wave (ref.) | --- | --- | --- | --- | --- | --- | --- | --- | --- | --- | --- | --- |
| Age 34 wave | **0.158** | **0.107, 0.209** | **0.159** | **0.108, 0.210** | **0.162** | **0.110, 0.215** | **0.153** | **0.115, 0.191** | **0.153** | **0.115, 0.191** | **0.157** | **0.118, 0.196** |
| Age 42 wave | **0.364** | **0.310, 0.419** | **0.369** | **0.315, 0.423** | **0.370** | **0.315, 0.426** | **0.340** | **0.299, 0.380** | **0.340** | **0.299, 0.380** | **0.343** | **0.302, 0.384** |
| Age 46/48 wave | **0.273** | **0.218, 0.329** | **0.280** | **0.224, 0.335** | **0.287** | **0.230, 0.344** | **0.261** | **0.220, 0.302** | **0.261** | **0.220, 0.302** | **0.262** | **0.221, 0.303** |
| **Interaction** | | | | | Joint $p = .733$ | | | | | | Joint $p = .730$ | |
| Veg. × Age 30 wave (ref.) |  |  |  |  |  |  |  |  |  |  |  |  |
| Veg. × Age 34 wave | --- | --- | --- | --- | -0.059 | -0.292, 0.174 | --- | --- | --- | --- | -0.099 | -0.289, 0.091 |
| Veg. × Age 42 wave | --- | --- | --- | --- | -0.021 | -0.262, 0.220 | --- | --- | --- | --- | -0.090 | -0.278, 0.098 |
| Veg. × Age 46/48 wave | --- | --- | --- | --- | -0.135 | -0.381, 0.111 | --- | --- | --- | --- | -0.033 | -0.227, 0.161 |

The outcome is time-varying psychological distress (0-9) between ages 30, 34, 42, and 46/48, defined through the 9-item Malaise Inventory. A higher score indicates more distress. Estimates are betas from random-intercept linear regression models. Control variables include: sex, intelligence at age 10, body mass index, self-rated health, and psychological distress at age 26, and educational attainment by age 30. Joint $p$ are based on Wald-type tests, computed with the Stata test and mi test commands. Bolded estimates are significant at the $p < .05$ level. CI = Confidence interval.

Before testing the interaction with time, the effect of being a vegetarian on psychological distress between ages 30 to 46/48 decreased from *b =* 0.279 (95%CI 0.090, 0.467) in the unadjusted model to *b =* 0.053 (95%CI -0.097, 0.203) in the fully-adjusted model, representing an 81.0% attenuation towards the null between models. The main effect of time shows that psychological distress significantly increased on average by 0.159 at age 34, 0.369 at age 42, and 0.280 at ages 46-48 compared to age 30. Testing the interaction with time, we found that vegetarianism was not associated with differences in within-person changes in psychological distress between ages 30 to 46/48 (*p* = .723).

### 3.4. Sensitivity analysis with red meat consumption

We reproduced the multilevel growth curve model with red meat consumption instead of self-reported vegetarianism using the same set of covariates and found similar results. Before testing the interaction with time, the effect of never eating red meat on psychological distress between ages 30 to 46/48 decreased from *b =* 0.235 (95%CI 0.092, 0.379) in the unadjusted model to *b =* 0.028 (95%CI -0.087, 0.144) in the fully-adjusted model, representing an 88.1% attenuation towards the null between models. Testing the interaction with time, we found that red meat consumption was not associated with differences in within-person changes in psychological distress between ages 30 to 46/48 (*p* = .251).

We report in Supplementary Figure 1 predicted scores based on models with the interaction for vegetarianism and red meat consumption, before and after statistical adjustment for covariates. Results show similar trends across the two indicators: those who identified as vegetarian and never ate red meat at age 30 reported on average a non-significant higher distress score at ages 30, 34, and 42, and a non-significant lower distress score at ages 46-48.

## 4. DISCUSSION

In response to the lack of robust evidence on the relationship between vegetarianism and mental health, we examined in a nationally representative sample within-person changes in psychological distress among adult vegetarians and non-vegetarians over a 16/18-year period and found no meaningful differences. While vegetarians reported more distress at age 30, this association was fully confounded by prior differences in mental distress and other factors such as sex, body mass index, and self-rated health. Using red meat consumption as a proxy yielded consistent findings.

The findings contribute to an emerging literature that nuances the argument that vegetarianism may be linked to mental health problems, and challenges the added value of new studies with weaker designs (e.g., cross-sectional design and poor statistical adjustment strategy). Supporting what Northstone et al. found among English parents in the mid-90s using ALSPAC data, we found that British vegetarians were not likely to have experienced meaningful differences in their mental health trajectory during adulthood compared to non-vegetarians.^22^ However, the fact that psychological distress at age 26 was a strong predictor of vegetarianism at age 30 (even when controlling for other covariates, see Supplementary Table 5) opens new questions about the nature of this relationship in earlier stages of the life-course. Young adulthood is an intense life stage in which identity development continues to occur.^38,39^ Vegetarianism may be part of this developmental process (and the stress involved), and no longer have a meaningful impact on mental health as one’s identity becomes entrenched in adulthood.^7^ Individuals also re-align behaviours including dietary habits in line with their values following a stressful event. Supporting this, one study found that people were likely to start vegetarianism following life transitions typically experienced in young adulthood such as moving, starting higher education, or relationship dissolution.^40^

We highlight that these findings may also not apply to other countries and newer generations. Vegetarianism is now relatively common in the United Kingdom, meaning that it may be easier here to start a vegetarian diet and maintain it over time compared with other Western countries. Vegetarianism may also be easier to maintain today compared with previous decades: e.g., environmental concerns are increasingly shared, plant-based options are easier to purchase in groceries and restaurants, and information on starting a vegetarian diet is easier to access. It may also have become easier to interact and develop a sense of belonging with other vegetarians over time through the use of information and communication technologies. That said, the lack of large surveys repeatedly collecting information on vegetarianism over time prevents us from knowing whether indicators such as changes in attitudes towards animal product consumption and spending on plant-based foods actually mean that there are more vegetarians. For instance, two studies in the UK and Finland have found that the number of vegetarian households has not meaningfully increased since the 1990s.^41,42^ Data in the US also suggests that there is instead an increasing proportion that self-identify as vegetarian yet eats some meat over time.^43^

### 4.1. Strengths and limitations

This study builds on the strengths of the 1970 British Cohort Study, which includes a nationally representative sample, frequent waves of follow-up, and relatively high response rates. Our findings are bound by the validity of our measure of vegetarianism and would be strengthened by more information on preference and behaviour (beyond red meat consumption). Using a single measurement of diet at age 30 also prevents us from considering if vegetarians changed diet during the follow-up period. Supporting the validity of our findings, the EPIC-Oxford study, a large cohort of UK adults started around the same time, found that vegetarians were highly likely to maintain their diet over time.^44^ Reproducing the analyses with other mental health measures such as subjective wellbeing, validated measures of depression and anxiety, and clinical diagnoses of mental health problems would also strengthen the validity of our findings. While this study benefitted from multiple measurements of psychological distress over time, the gold standard for causal inference in observational studies includes having multiple measurements of the exposure and considering within-person change in the exposure and the outcome (i.e., what is the effect of *becoming* and *no longer being* vegetarian on mental health).

### 4.2 Conclusion

This is one of the more robust studies to have investigated the relationship between vegetarianism and mental health. Supporting some of the stronger studies published to date, we did not find a substantive relationship between vegetarianism and psychological distress during adulthood in this cohort representative of those born across Great Britain in 1970. Their association across earlier life stages, however, is still unclear. The relationship between diet and mental health is complex and includes both biological, psychological, and social mechanisms. Studies seeking to shed new light on these issues require both strong data, methods, and theory.

## Data Availability

Data analysed in this study are available online on the UK Data Service platform.

https://beta.ukdataservice.ac.uk/datacatalogue/studies/study?id=8547#!/access-data

## ACKNOWLEDGEMENTS

VK was affiliated with the University of Reading during the writing of this project. Data from the 1970 British Cohort Study was accessed on the UK Data Service platform. Reference: University of London, Institute of Education, Centre for Longitudinal Studies. (2021). 1970 British Cohort Study: Age 46, Sweep 10, 2016-2018. [data collection]. UK Data Service. SN: 8547, DOI: 10.5255/UKDA-SN-8547-1

## AUTHOR CONTRIBUTIONS

**TG:** Conceptualisation; Methodology; Formal analysis; Writing – Original Draft; Writing – Review & Editing. **VK:** Writing – Review & Editing.

## ETHICAL STATEMENT

There was no need to obtain ethical approval for the secondary data analysis of this dataset. Detailed information on ethical review and consent in the 1970 British Cohort Study is outlined elsewhere (Shepherd and Gilbert, 2019).

Shepherd P, Gilbert E. 2019. 1970 British Cohort Study. Ethical review and consent. URL: https://cls.ucl.ac.uk/wp-content/uploads/2017/07/BCS70-Ethical-review-and-Consent-2019.pdf

## REFERENCES

1. Dinu M, Abbate R, Gensini GF, Casini A, Sofi F. Vegetarian, vegan diets and multiple health outcomes: A systematic review with meta-analysis of observational studies. Crit Rev Food Sci Nutr. 2017;57(17):3640–3649. doi:10.1080/10408398.2016.1138447

2. Poore J, Nemecek T. Reducing food’s environmental impacts through producers and consumers. Science. 2018;360(6392):987–992. doi:10.1126/science.aaq0216

3. Kim B, Neff R, Santo R, Vigorito J. The Importance of Reducing Animal Product Consumption and Wasted Food in Mitigating Catastrophic Climate Change. Published online 2015. https://clf.jhsph.edu/sites/default/files/2019-01/importance-of-reducing-animal-product-consumption-and-wasted-food-in-mitigating-catastrophic-climate-change.pdf

4. Orlich MJ, Chiu THT, Dhillon PK, et al. Vegetarian Epidemiology: Review and Discussion of Findings from Geographically Diverse Cohorts. Adv Nutr. 2019;10(Supplement_4):S284–S295. doi:10.1093/advances/nmy109

5. Li F, Liu X, Zhang D. Fish consumption and risk of depression: a meta-analysis. J Epidemiol Community Health. 2016;70(3):299–304. doi:10.1136/jech-2015-206278

6. Hargreaves SM, Raposo A, Saraiva A, Zandonadi RP. Vegetarian Diet: An Overview through the Perspective of Quality of Life Domains. Int J Environ Res Public Health. 2021;18(8):4067. doi:10.3390/ijerph18084067

7. Dl R, Al B. The unified model of vegetarian identity: A conceptual framework for understanding plant-based food choices. Appetite. 2017;112. doi:10.1016/j.appet.2017.01.017

8. Dobersek U, Teel K, Altmeyer S, Adkins J, Wy G, Peak J. Meat and mental health: A meta-analysis of meat consumption, depression, and anxiety. Crit Rev Food Sci Nutr. 2021;0(0):1–18. doi:10.1080/10408398.2021.1974336

9. Iguacel I, Huybrechts I, Moreno LA, Michels N. Vegetarianism and veganism compared with mental health and cognitive outcomes: a systematic review and meta-analysis. Nutr Rev. 2021;79(4):361–381. doi:10.1093/nutrit/nuaa030

10. Perry CL, Mcguire MT, Neumark-Sztainer D, Story M. Characteristics of vegetarian adolescents in a multiethnic urban population1 1The full text of this article is available via JAH Online at http://www.elsevier.com/locate/jahonline. J Adolesc Health. 2001;29(6):406–416. doi:10.1016/S1054-139X(01)00258-0

11. Baines S, Powers J, Brown WJ. How does the health and well-being of young Australian vegetarian and semi-vegetarian women compare with non-vegetarians? Public Health Nutr. 2007;10(5):436–442. doi:10.1017/S1368980007217938

12. Dobersek U, Wy G, Adkins J, et al. Meat and mental health: a systematic review of meat abstention and depression, anxiety, and related phenomena. Crit Rev Food Sci Nutr. 2021;61(4):622–635. doi:10.1080/10408398.2020.1741505

13. Askari M, Daneshzad E, Darooghegi Mofrad M, Bellissimo N, Suitor K, Azadbakht L. Vegetarian diet and the risk of depression, anxiety, and stress symptoms: a systematic review and meta-analysis of observational studies. Crit Rev Food Sci Nutr. 2020;0(0):1–11. doi:10.1080/10408398.2020.1814991

14. Kris-Etherton PM, Petersen KS, Hibbeln JR, et al. Nutrition and behavioral health disorders: depression and anxiety. Nutr Rev. 2020;79(3):247–260. doi:10.1093/nutrit/nuaa025

15. Medawar E, Huhn S, Villringer A, Veronica Witte A. The effects of plant-based diets on the body and the brain: a systematic review. Transl Psychiatry. 2019;9:226. doi:10.1038/s41398-019-0552-0

16. Paslakis G, Richardson C, Nöhre M, et al. Prevalence and psychopathology of vegetarians and vegans – Results from a representative survey in Germany. Sci Rep. 2020;10:6840. doi:10.1038/s41598-020-63910-y

17. Rossa-Roccor V, Richardson CG, Murphy RA, Gadermann AM. The association between diet and mental health and wellbeing in young adults within a biopsychosocial framework. PLoS ONE. 2021;16(6):e0252358. doi:10.1371/journal.pone.0252358

18. Hibbeln JR, Northstone K, Evans J, Golding J. Vegetarian diets and depressive symptoms among men. J Affect Disord. 2018;225:13–17. doi:10.1016/j.jad.2017.07.051

19. Michalak J, Zhang XC, Jacobi F. Vegetarian diet and mental disorders: results from a representative community survey. Int J Behav Nutr Phys Act. 2012;9:67. doi:10.1186/1479-5868-9-67

20. Matta J, Czernichow S, Kesse-Guyot E, et al. Depressive Symptoms and Vegetarian Diets: Results from the Constances Cohort. Nutrients. 2018;10(11):1695. doi:10.3390/nu10111695

21. Schreiner P, Yilmaz B, Rossel JB, et al. Vegetarian or gluten-free diets in patients with inflammatory bowel disease are associated with lower psychological well-being and a different gut microbiota, but no beneficial effects on the course of the disease. United Eur Gastroenterol J. 2019;7(6):767–781. doi:10.1177/2050640619841249

22. Northstone K, Joinson C, Emmett P. Dietary patterns and depressive symptoms in a UK cohort of men and women: a longitudinal study. Public Health Nutr. 2018;21(5):831–837. doi:10.1017/S1368980017002324

23. Velten J, Bieda A, Scholten S, Wannemüller A, Margraf J. Lifestyle choices and mental health: a longitudinal survey with German and Chinese students. BMC Public Health. 2018;18(1):632. doi:10.1186/s12889-018-5526-2

24. Lavallee K, Zhang XC, Michalak J, Schneider S, Margraf J. Vegetarian diet and mental health: Cross-sectional and longitudinal analyses in culturally diverse samples. J Affect Disord. 2019;248:147–154. doi:10.1016/j.jad.2019.01.035

25. Shen YC, Chang CE, Lin MN, Lin CL. Vegetarian Diet Is Associated with Lower Risk of Depression in Taiwan. Nutrients. 2021;13(4):1059. doi:10.3390/nu13041059

26. Pitchforth J, Fahy K, Ford T, Wolpert M, Viner RM, Hargreaves DS. Mental health and well-being trends among children and young people in the UK, 1995-2014: analysis of repeated cross-sectional national health surveys. Psychol Med. 2019;49(8):1275–1285. doi:10.1017/S0033291718001757

27. Molendijk M, Molero P, Ortuño Sánchez-Pedreño F, Van der Does W, Angel Martínez-González M. Diet quality and depression risk: A systematic review and dose-response meta-analysis of prospective studies. J Affect Disord. 2018;226:346–354. doi:10.1016/j.jad.2017.09.022

28. Gale CR, Deary IJ, Schoon I, Batty GD. IQ in childhood and vegetarianism in adulthood: 1970 British cohort study. BMJ. 2007;334(7587):245. doi:10.1136/bmj.39030.675069.55

29. Rodgers B, Pickles A, Power C, Collishaw S, Maughan B. Validity of the Malaise Inventory in general population samples. Soc Psychiatry Psychiatr Epidemiol. 1999;34(6):333–341. doi:10.1007/s001270050153

30. Ploubidis GB, Sullivan A, Brown M, Goodman A. Psychological distress in mid-life: evidence from the 1958 and 1970 British birth cohorts. Psychol Med. 2017;47(2):291–303. doi:10.1017/S0033291716002464

31. Gondek D, Bann D, Patalay P, et al. Psychological distress from early adulthood to early old age: evidence from the 1946, 1958 and 1970 British birth cohorts. Psychol Med. Published online January 21, 2021:1–10. doi:10.1017/S003329172000327X

32. Parsons S. Childhood cognition in the 1970 British Cohort Study. Published online 2014.

33. Curran PJ, Obeidat K, Losardo D. Twelve Frequently Asked Questions About Growth Curve Modeling. J Cogn Dev Off J Cogn Dev Soc. 2010;11(2):121–136. doi:10.1080/15248371003699969

34. Williams R. Using the Margins Command to Estimate and Interpret Adjusted Predictions and Marginal Effects. Stata J. 2012;12(2):308–331. doi:10.1177/1536867X1201200209

35. Statacorp. Stata Statistical Software: Release 16. Published online 2019.

36. Royston P, White IR. Multiple Imputation by Chained Equations (MICE): Implementation in Stata. J Stat Softw. 2011;45(1):1–20. doi:10.18637/jss.v045.i04

37. Mostafa T, Wiggins R. The impact of attrition and non-response in birth cohort studies: a need to incorporate missingness strategies. Longitud Life Course Stud. 2015;6(2):131–146. doi:10.14301/llcs.v6i2.312

38. Arnett JJ. Emerging adulthood. A theory of development from the late teens through the twenties. Am Psychol. 2000;55(5):469–480.

39. Côté JE. Sociological perspectives on identity formation: the culture–identity link and identity capital. J Adolesc. 1996;19(5):417–428. doi:10.1006/jado.1996.0040

40. Jabs J, Devine CM, Sobal J. Model of the Process of Adopting Vegetarian Diets: Health Vegetarians and Ethical Vegetarians. J Nutr Educ. 1998;30(4):196–202. doi:10.1016/S0022-3182(98)70319-X

41. Waters J. A model of the dynamics of household vegetarian and vegan rates in the U.K. Appetite. 2018;127:364–372. doi:10.1016/j.appet.2018.05.017

42. Vinnari M, Mustonen P, Räsänen P. Tracking down trends in non-meat consumption in Finnish households, 1966-2006. Br Food J. 2010;112(8):836–852. doi:10.1108/00070701011067451

43. Šimčikas S. Is the Percentage of Vegetarians and Vegans in the U.S. Increasing? Published August 16, 2018. Accessed October 29, 2021. https://animalcharityevaluators.org/blog/is-the-percentage-of-vegetarians-and-vegans-in-the-u-s-increasing/

44. Key TJ, Appleby PN, Spencer EA, Travis RC, Roddam AW, Allen NE. Mortality in British vegetarians: results from the European Prospective Investigation into Cancer and Nutrition (EPIC-Oxford). Am J Clin Nutr. 2009;89(5):1613S–1619S. doi:10.3945/ajcn.2009.26736L

