## Supplementary Material for "Vegetarianism and mental health: longitudinal evidence in the 1970 British Cohort Study"

**TITLE**

**TABLE OF CONTENTS**

**1. TABLE S1. Response rates in the 1970 British Cohort Study.**

**2. TABLE S2. Missing data on variables in the 1970 British Cohort Study.**

**3. TABLE S3. Differences in distress by red meat (RM) consumption at age 30.**

**4. TABLE S4. Differences in distress by red meat (RM) consumption between ages 30 and 46/48.**

**5. FIGURE S1. Trajectories of psychological distress by red meat (RM) consumption status.**

**6. TABLE S5. Predicting vegetarianism at age 30.**

**SUPPLEMENTARY TABLE 1**

**Response rates among eligible cases in the 1970 British Cohort Study (1970 to 2016/18).**

|  | **Response rate among all eligible cases** | **Response rate among productive cases at age 30** |
| --- | --- | --- |
|  | **%** | **%** |
| **Age 10 wave** | 90.4^1^ | 92.5 |
| **Age 26 wave** | 59.8^1^ | 67.8 |
| **Age 30 wave** | 74.7^1^ | --- |
| **Age 34 wave** | 75.0^2^ | 79.9 |
| **Age 42 wave** | 74.6^3^ | 74.9 |
| **Age 46/48 wave** | 73.0^4^ | 65.7 |

^1^ These response rates, which refer to cohort members surveyed at birth and other cohort members included over the course of the study in later waves, were estimated by Plewis et al. (2004).

Plewis I, Calderwood L, Hawkes D, Nathan G. 2004. Changes in the NCDS and BCS70 Populations and Samples over Time, First edition. Center for Longitudinal Studies, University College London.

^2^ URL: https://cls.ucl.ac.uk/wp-content/uploads/2017/06/0-Technical-report-Final-16082007.pdf

^3^ URL: https://cls.ucl.ac.uk/wp-content/uploads/2017/07/BCS70-Technical-report-FINAL.pdf

^4^ URL: https://cls.ucl.ac.uk/wp-content/uploads/2019/03/BCS46_Technical-Report_FINAL.pdf

**SUPPLEMENTARY TABLE 2**

**Missing data on variables in the 1970 British Cohort Study (1970 to 2016/18).**

|  | **Productive cases at age 30** |
| --- | --- |
|  | ***N* = 11,216** |
|  | **%** |
| Vegetarianism at age 30 | 0.5 |
| Red meat consumption at age 30 | 0.5 |
| Distress at age 30 | 1.4 |
| Distress at age 34 | 20.7 |
| Distress at age 42 | 35.1 |
| Distress at age 46/48 | 39.9 |
| **Age 10** |  |
| Sex | 0.0 |
| Intelligence | 28.9 |
| Physical/mental limitations | 14.4 |
| Family social class | 8.0 |
| Country | 8.3 |
| Mother’s meat consumption | 21.7 |
| **Age 26** |  |
| Cohabitation | 33.1 |
| Living with children | 33.1 |
| Economic activity | 32.9 |
| Education * | 0.3 |
| Body mass index * | 33.8 |
| Self-rated health | 32.5 |
| Life satisfaction | 32.6 |
| Distress | 34.0 |

* These variables were coded based on information in the age 30 wave.

**SUPPLEMENTARY TABLE 3**

**Differences in psychological distress by red meat consumption status at age 30 in the 1970 British Cohort Study (1970 to 2016/18).**

|  | **Complete-case sample** | | | **Multiple imputation** | | |
| --- | --- | --- | --- | --- | --- | --- |
|  | ***n* = 4,658** | | | ***n* = 11,261** | | |
|  | **B** | **95%CI** | **% reduced** | **B** | **95%CI** | **% reduced** |
| **Never eats red meat (ref. = Yes)** | **0.292** | **0.130, 0.454** | **---** | **0.254** | **(0.138, 0.371)** | **---** |
| **Controlling for …** |  |  |  |  |  |  |
| **At age 10** |  |  |  |  |  |  |
| Sex | **0.181** | **0.019, 0.343** | 38.0 | **0.142** | **(0.024, 0.260)** | 44.1 |
| Intelligence | **0.324** | **0.163, 0.485** | -11.0 | **0.286** | **(0.170, 0.402)** | -12.6 |
| Health limitations | **0.292** | **0.131, 0.454** | 0.0 | **0.260** | **(0.144, 0.377)** | -2.4 |
| Family social class | **0.296** | **0.134, 0.457** | -1.4 | **0.258** | **(0.142, 0.374)** | -1.6 |
| Country | **0.293** | **0.131, 0.456** | -0.3 | **0.255** | **(0.139, 0.372)** | -0.4 |
| Mother’s meat consumption | **0.271** | **0.108, 0.434** | 7.2 | **0.236** | **(0.119, 0.353)** | 7.1 |
| **At age 26** |  |  |  |  |  |  |
| Cohabitation | **0.281** | **0.118, 0.443** | 3.8 | **0.244** | **(0.126, 0.361)** | 3.9 |
| Living with children | **0.305** | **0.143, 0.467** | -4.5 | **0.276** | **(0.160, 0.393)** | -8.7 |
| Economic activity | **0.257** | **0.094, 0.420** | 12.0 | **0.218** | **(0.100, 0.336)** | 14.2 |
| Education | **0.331** | **0.168, 0.492** | -13.4 | **0.308** | **(0.192, 0.425)** | -21.3 |
| Body mass index | **0.258** | **0.096, 0.420** | 11.6 | **0.230** | **(0.113, 0.347)** | 9.4 |
| Self-rated health | **0.263** | **0.102, 0.424** | 9.9 | **0.220** | **(0.105, 0.336)** | 13.4 |
| Life satisfaction | **0.288** | **0.128, 0.449** | 1.4 | **0.259** | **(0.143, 0.375)** | -2.0 |
| Psychological distress at ages 26 | 0.076 | -0.066, 0.219 | 74.0 | 0.057 | (-0.051, 0.165) | 77.6 |
| **All 14 variables at ages 10 and 26** | 0.021 | -0.123, 0.165 | 92.8 | 0.016 | (-0.092, 0.125) | 93.7 |

The outcome is psychological distress (0-9) at age 30, defined through the 9-item Malaise Inventory. A higher score indicates higher distress. Estimates represent mean differences in the outcome between never red meat eaters and others, controlling for other variables. The “% reduced” represents the attenuation by statistical adjustment and is computed from the ratio of estimates (e.g., 1 - (0.021/0.292) = 92.8%). Bolded estimates are significant at the *p* < .05 level. CI = Confidence interval.

**SUPPLEMENTARY TABLE 4**

**Differences in psychological distress by red meat consumption status between ages 30 and 46/48 in the 1970 British Cohort Study (1970 to 2016/18).**

|  | **Complete-case sample** | | | | | | **Multiple imputation (20 datasets)** | | | | | |
| --- | --- | --- | --- | --- | --- | --- | --- | --- | --- | --- | --- | --- |
|  | ***N* participants = 4,909; *N* observations = 16,727** | | | | | | ***N* participants = 11,261; *n* observations *=* 45,044** | | | | | |
|  | **No controls** | | **With controls** | | **With interaction** | | **No controls** | | **With controls** | | **With interaction** | |
|  | **B** | **95%CI** | **B** | **95%CI** | **B** | **95%CI** | **B** | **95%CI** | **B** | **95%CI** | **B** | **95%CI** |
| **Never eating red meat (ref. = No)** | **0.235** | **0.092, 0.379** | 0.028 | -0.087, 0.144 | 0.089 | -0.063, 0.241 | **0.221** | **0.112, 0.331** | 0.039 | -0.061, 0.139 | 0.072 | -0.045, .189 |
| **Time** |  |  |  |  |  |  |  |  |  |  |  |  |
| Age 30 wave (ref.) | --- | --- | --- | --- | --- | --- | --- | --- | --- | --- | --- | --- |
| Age 34 wave | **0.158** | **0.107, 0.210** | **0.159** | **0.108, 0.210** | **0.161** | **0.107, 0.214** | **0.153** | **0.115, 0.191** | **0.153** | **0.115, 0.191** | **0.155** | **0.112, 0.191** |
| Age 42 wave | **0.365** | **0.310, 0.419** | **0.369** | **0.315, 0.423** | **0.379** | **0.322, 0.436** | **0.340** | **0.300, 0.380** | **0.340** | **0.300, 0.380** | **0.347** | **0.306, 0.389** |
| Age 46/48 wave | **0.273** | **0.218, 0.329** | **0.280** | **0.224, 0.335** | **0.296** | **0.238, 0.354** | **0.261** | **0.220, 0.302** | **0.261** | **0.220, 0.302** | **0.267** | **0.223, 0.310** |
| **Interaction** |  |  |  | | Joint p = .251 | |  |  |  | | Joint p = .479 | |
| Veg. × Age 30 wave (ref.) | --- | --- | --- | --- | --- | --- |  |  |  |  | --- | --- |
| Veg. × Age 34 wave | --- | --- | --- | --- | -0.014 | -0.188, 0.160 |  |  |  |  | -0.013 | -0.125, 0.151 |
| Veg. × Age 42 wave | --- | --- | --- | --- | -0.099 | -0.282, 0.083 |  |  |  |  | -0.084 | -0.223, 0.056 |
| Veg. × Age 46/48 wave | --- | --- | --- | --- | -0.173 | -0.361, 0.014 |  |  |  |  | -0.062 | -0.199, 0.074 |

The outcome is time-varying psychological distress (0-9) between ages 30, 34, 42, and 46/48, defined through the 9-item Malaise Inventory. Estimates are betas from random-intercept linear regression models. Control variables include: sex, intelligence at age 10, body mass index, self-rated health, and psychological distress at age 26, and educational attainment by age 30. Joint *p* are based on Wald-type tests, computed with the Stata *test* and *mi test* commands. Bolded estimates are significant at the *p* < .05 level. CI = Confidence interval.

**SUPPLEMENTARY FIGURE 1**

**Trajectories of psychological distress, before and after statistical adjustment, in the 1970 British Cohort Study (1970 to 2016/18).**

Panel B: Red meat consumption at age 30

|  | Unadjusted Red meat | Unadjusted Never red meat | Fully-adjusted Red meat | Fully-adjusted Never red meat |
| --- | --- | --- | --- | --- |
| Age 30 | 1.41549 | 1.70806 | 1.42001 | 1.50916 |
| Age 34 | 1.57484 | 1.85577 | 1.58061 | 1.65538 |
| Age 42 | 1.78982 | 1.9828 | 1.79877 | 1.78897 |
| Ages 46-48 | 1.70446 | 1.83433 | 1.71626 | 1.63196 |

Panel A: Vegetarianism at age 30

Psychological distress score (0-9)

|  | Unadjusted Not vegetarian | Unadjusted Vegetarian | Fully-adjusted Not vegetarian | Fully-adjusted Vegetarian |
| --- | --- | --- | --- | --- |
| Age 30 | 1.42789 | 1.7382 | 1.4235 | 1.52361 |
| Age 34 | 1.58893 | 1.84215 | 1.58569 | 1.62678 |
| Age 42 | 1.79306 | 2.08872 | 1.79367 | 1.87281 |
| Ages 46-48 | 1.70755 | 1.89413 | 1.71025 | 1.67569 |

Estimates are adjusted average marginal means based on the “complete-case” models in Table 3 and Supplementary Table 4.

A higher score indicates higher distress.

**SUPPLEMENTARY TABLE 5**

**Predicting vegetarianism at age 30 in the** **1970 British Cohort Study (2000).**

|  | **Complete case sample (n *=* 4,685)** | | | | | | **Multiple imputation (*n* = 11,261)** | | | | | |
| --- | --- | --- | --- | --- | --- | --- | --- | --- | --- | --- | --- | --- |
|  | **Bivariate** | | **+ Age 10 controls** | | **+ Age 26 controls** | | **Bivariate** | | **+ Age 10 controls** | | **+ Age 26 controls** | |
|  | **PR** | **95%CI** | **PR** | **95%CI** | **PR** | **95%CI** | **PR** | **95%CI** | **PR** | **95%CI** | **PR** | **95%CI** |
| **Psychol. distress at age 26** | **1.12** | **1.06-1.19** | **1.09** | **1.03-1.16** | **1.09** | **1.02-1.17** | **1.11** | **1.06-1.16** | **1.09** | **1.04-1.14** | **1.09** | **1.04-1.14** |

Estimates are prevalence ratios (PR) from Poisson models with robust variance estimation. Estimates represent the added relative probability of identifying as vegetarian for a one-unit increase in the psychological distress score (range: 0-9) at age 26. Bolded estimates are significant at the .05 level.

Age 10 controls include: sex, intelligence, health limitations, family social class, mother’s meat consumption, and country of residence. Age 26 controls also include: education, cohabitation, economic activity, parenthood, life satisfaction, and self-rated health.
